# Association between the *ACE* I/D gene polymorphism and progressive renal failure in autosomal dominant polycystic kidney disease: A meta-analysis

**DOI:** 10.1101/19002949

**Authors:** Noel Pabalan, Phuntila Tharabenjasin, Yardnapar Parcharoen, Adis Tasanarong

## Abstract

**Objective:** The *angiotensin converting enzyme* insertion/deletion (*ACE* I/D) gene polymorphism is involved in a wide range of clinical outcomes. This makes *ACE* I/D an important genetic marker. Updating the genetic profile of *ACE* I/D and raising the evidence for its role in renal disease is therefore needed. Reported associations of *ACE* I/D with progressive renal failure (PRF) in autosomal dominant polycystic kidney disease (ADPKD) have been inconsistent, prompting a meta-analysis to obtain more precise estimates.

**Methods:** Multi-database search yielded 18 articles for inclusion in the meta-analysis. Risks (odds ratios [ORs] and 95% confidence intervals) were estimated by comparing the *ACE* genotypes (heterozygote ID, homozygotes DD and II). Heterogeneous (random-effects) pooled associations were subjected to outlier treatment which yielded fixed-effects outcomes and split the findings into pre- (PRO) and post- (PSO) outlier status. Subgroup analysis was based on ethnicity (Asian/Caucasian) and minor allele frequency (maf). The ≥ 0.50 maf subgroup indicates higher frequency of the variant II genotype over that of the common DD genotype, otherwise, the subgroup is considered < 0.50 maf. Stability of the associative effects was assessed with sensitivity treatment. Temporal trend of association was examined with cumulative meta-analysis.

**Results:** In the PSO analysis, overall effects were null (ORs 0.99-1.02) but not in the subgroups (Asian and ≥ 0.50 maf), where in presence of the D allele (DD/ID) and the I allele (II), increased (ORs 1.63-5.62) and reduced (OR 0.22) risks were observed, respectively. Of these pooled effects, the Asian and ≥ 0.50 maf homozygous DD genotypes had high ORs (5.01-5.63) indicating elevated magnitude of effects that were highly significant (P^a^ < 10^−5^) and homogeneous (I^*2*^ = 0%), in addition to their robustness. In contrast, the Caucasian and < 0.50 maf subgroup effects were: (i) non-heterogeneous (fixed-effects) at the outset, which did not require outlier treatment and (ii) non-significant (ORs 0.91-1.10, P^a^ = 0.15-0.79). Cumulative meta-analysis revealed increased precision of effects over time.

**Conclusions:** PRF in ADPKD impacted the Asian and ≥ 0.50 maf subgroups where DD homozygote carriers were up to 6-fold susceptible. The high magnitude of these effects were highly significant, homogeneous and robust indicating strong evidence of association.

## Introduction

Autosomal dominant polycystic kidney disease (ADPKD) is an inherited systemic disease characterized by fluid filled cysts in the kidneys leading to end stage renal failure in later years of life [1]. Preventing progressive renal failure (PRF) is mainly treated with controlling blood pressure and reducing proteinuria [2]. However, these reno-protective measures do not benefit all patients [3]. These differential treatment outcomes may in part be attributed to genetics. Studies on mouse and human models suggest that genetic variation plays a significant role in PRF associated with ADPKD [4,5]. One genetic variation is the insertion/ deletion (I/D) polymorphism in the *angiotensin converting enzyme* (*ACE*) gene, a 24 kb sequence of DNA in intron 16 of chromosome 17q23. This gene encodes the ACE protein, which is found in mammalian tissues and body fluids as ectoenzymes on cell surfaces and in serum [6]. The I/D polymorphism has been shown to determine the levels of circulating ACE enzymes [7]. ACE enzyme levels have been suggested to associate with the *ACE* genotypes (heterozygote ID, homozygotes DD and II) possibly affecting therapeutic response [8].

Individuals carrying the DD homozygote genotype were found to exhibit the highest serum ACE activity when compared to carriers of II homozygote and ID heterozygote genotype who showed low and intermediate activity, respectively [7]. The relationship between ACE activity and *ACE* genotypes, specifically homozygous DD appears to be the central concept in the clinical genetics of renal disease [9]. In a modification of ACE activity, a meta-analysis of 46 studies revealed that the genotypes (ID, DD) containing the D allele showed higher plasma ACE activity than the homozygous II genotype [10]. In ADPKD patients, the DD homozygote genotype was found to correlate with progression of renal insufficiency [11]. In addition, ADPKD patients have hypertension, a severe complication in which the *ACE* gene is most likely involved [10]. The value of *ACE* I/D as a genetic marker lies in its association with risk of a wide range of clinical outcomes that include response to ACE inhibitor therapy [12], cardiovascular diseases [10], diabetes-related [13,14], cancer [15], longevity [16], Alzheimer’s disease [17] and muscle performance [18]. The span of *ACE* involvement in a wide variety of clinical conditions and evidence of elevated ACE activity associated with the *ACE* D allele prompted a Genome-Wide Association Study (GWAS) from the quantitative trait loci perspective [19], with no mention of ADPKD. Another GWAS profiled ADPKD from an epigenetic perspective [20], with no mention of *ACE* I/D. Thus, GWAS has involved *ACE* I/D and ADPKD which were not reported in the same study. One study that examined both *ACE* I/D and ADPKD was a meta-analysis, which was published in 2006 [21]. Since then, new primary studies have emerged, with inconsistent results. Given the variability of results and length of time (13 years) since the last synthesis, we undertook this meta-analysis for three reasons: (i) obtain less ambiguous, clearer estimates and updated role of *ACE* I/D with renal failure progression in ADPKD; (ii) apply novel meta-analysis techniques (e.g. outlier treatment) in order to raise the strength of evidence and (iii) examine the cumulative trend of association. Simultaneous application of meta-analysis treatments that focus on the pathophysiological role of *ACE* I/D precluded inclusion of other polymorphisms in this study. This meta-analysis aims for better understanding of the genetics of PRF in ADPKD, so that it may provide important information that might be useful to decision makers in healthcare, particularly in the field of nephrology.

## Materials and Methods

### Selection of studies

Two authors (NP and PT) performed primary screening (titles/abstracts) and disagreements were resolved through screening of the title/abstract in question by a third author (YP). Three databases (PubMed in MEDLINE, Google Scholar and Science Direct) were searched for association studies as of March 12, 2019. Terms used were “*angiotensin converting enzyme”*, “*ACE*”, “ADPKD”, “autosomal dominant polycystic kidney disease” and “polymorphism” as medical subject heading and text, restricted to the English language. Additional eligible studies were identified from references cited in the retrieved articles. Inclusion criteria were: (i) case–control study design evaluating the association between *ACE* I/D and PRF in ADPKD and (ii) sufficient genotype or allele frequency data to allow calculation of odds ratios (ORs) and 95% confidence intervals (CIs). Exclusion criteria are: (i) studies without controls or studies whose genotype or allele frequencies were unusable/absent; (ii) those that did not cover the polymorphism or disease in question, (iii) reviews and (iv) non-English articles (S1 List).

**S1 List** Excluded studies

### Data extraction

Two investigators independently extracted the data that resulted in consensus. Extracted information from each article included the first author’s name, publication year, country of origin, ethnicity, study design and whether the articles addressed the Hardy-Weinberg Equilibrium (HWE). Articles which were not included in a previous meta-analysis [21] are indicated by an asterisk under the author column (Table 1). Primary study authors were contacted in order to obtain more information on incomplete data. Less than a third of the included studies mentioned influence of the environment, but data were not provided.

**Table 1.** Characteristics of the included studies that examined *ACE I/D* polymorphism associations with PRF in ADPKD

|  | First author | [R] | Year | Country | Ethnicity | Study design | Addressed HWE | CB |
| --- | --- | --- | --- | --- | --- | --- | --- | --- |
| 1 | Konoshita | [24] | 2001 | Japan | Asian | CS | Yes | 7 |
| 2 | Lee | [25] | 2000 | Korea | Asian | CS | NM | 5 |
| 3 | Ramanathan * | [26] | 2016 | India | Asian | CC | Yes | 6 |
| 4 | Tripathi | [27] | 2006 | India | Asian | CC** | NM | 7 |
| 5 | Tripathi * | [28] | 2008 | India | Asian | CC** | NM | 8 |
| 6 | Uemasu | [29] | 1997 | Japan | Asian | CC | NM | 5 |
| 7 | Baboolal | [30] | 1997 | UKA | Caucasian <sup>a</sup> | CS | Yes | 7 |
| 8 | Buraczynska * | [31] | 2006 | Poland | Caucasian | CC** | Yes | 7 |
| 9 | Ecder | [32] | 2003 | USA | Caucasian | CS | Yes | 8 |
| 10 | Gumprecht * | [33] | 2007 | Poland | Caucasian | CS | NM | 4 |
| 11 | Lovati * | [34] | 2001 | Switzerland | Caucasian <sup>a</sup> | CC** | Yes | 9 |
| 12 | Merta | [35] | 2003 | Czechoslovakia | Caucasian | CC | Yes | 5 |
| 13 | Perez-Oller | [36] | 1999 | Spain- | Caucasian | CS | NM | 6 |
| 14 | Persu | [37] | 2003 | BFR | Caucasian | CS | Yes | 6 |
| 15 | Saggar-Malik | [38] | 2000 | UK | Caucasian | CO | NM | 6 |
| 16 | Schiavello | [39] | 2001 | ABP | Caucasian | CS | Yes | 6 |
| 17 | van Dijk * | [40] | 2000 | Netherlands | Caucasian | CC** | Yes | 6 |
| 18 | Wanic-Kossowska * | [41] | 2014 | Poland | Caucasian | CC** | NM | 7 |

*ACE*: angiotensin converting enzyme gene; *I/D*: polymorphism; PRF: progressive renal failure; ADPKD: autosomal dominant polycystic kidney disease; [R]: reference number of the studies; UKA: United Kingdom Australia; USA: United States of America; BFR: Belgium France; ABP: Australia Bulgaria Poland; CS: cross-sectional; CC: case-control; CO: cohort; \*\* diluted cases (ADPKD was one of the disease conditions of the patients); HWE: Hardy-Weinberg Equilibrium; NM: not mentioned; CB: Clark-Baudouin; \*not in the Pereira (2006) meta-analysis; Caucasian <sup>a</sup>: admixed population

### Data distribution, power calculations and HWE assessment

Data distribution was assessed with the Shapiro-Wilks (SW) test using SPSS 20.0 (IBM Corp., Armonk, NY, USA). Normal distribution (P > 0.05) warranted descriptive and inferential expressions of mean ± standard deviation (SD) and the parametric approach, respectively. Otherwise, the median (with interquartile range) and non-parametric tests were used, respectively. Using the G*Power program [22], we evaluated statistical power as its adequacy bolsters the level of associative evidence. Assuming an OR of 1.5 at a genotypic risk level of α = 0.05 (two-sided), power was considered adequate at ≥ 80%. HWE was assessed using the application in https://ihg.gsf.de/cgi-bin/hw/hwa1.pl.

### Methodological quality of the studies

We used the Clark-Baudouin (CB) scale to evaluate methodological quality of the included studies [23]. The CB criteria include P-values, statistical power, correction for multiplicity, comparative sample sizes between cases and controls, genotyping methods and the HWE. In this scale, low, moderate and high have scores of < 5, 5-6 and ≥ 7, respectively.

### Genetic models

From the 18 included studies [24-41], minor allele frequency (maf) of the homozygous II genotype among controls was < 0.50 in 11 of them [25,30-33,35-37,39-41] and not (≥ 0.50) the remaining seven [24,26-29,34,38]. Non-uniformity of the maf across the studies (S1 Table) influenced our study design in two ways: (1) maf data were dichotomized into ≥ 0.50 maf (DD < II) and < 0.50 maf (DD > II) and (2) precluded the use of standard genetic modeling. Thus, we opted for the allele-genotype model, wherein we compared (i) DD homozygotes with the ID/II genotype; (ii) II homozygotes with ID/DD genotype and (iii) heterozygous ID genotype carriers with homozygotes of the II and DD genotypes.

### Data synthesis

Risks of PRF in ADPKD (using raw data for frequencies) were estimated for each study and comparing the effects on the same baseline, we calculated pooled ORs. We performed a cumulative meta-analysis to examine changes in the pooled ORs with accumulation of data over time. Subgrouping was ethnicity-based (Asians and Caucasians) and maf-based (≥ 0.50 maf and < 0.50 maf). To assess the strength of evidence, we used three indicators: First, the magnitude of effects are higher or lower when the pooled ORs are farther from or closer to the OR value of 1.0 (null effect), respectively [42]. Second, the P-value is contextualized from the Bayesian perspective of the Bayes Factor (BF). The BF compares support from the null and alternate hypotheses, in contrast to the P-value, which addresses the null only [43]. Thus, P-values (Z-scores) of 0.05 and 0.001 (corresponding to minimum BFs of ≥ 0.15 and 0.005) indicate moderate and strong (to very strong) evidence, respectively [44]. The BF rests on the likelihood paradigm [45], indicative of how strong the hypotheses are supported by the data [43]. Thus, the likelihoods between the absence (null hypothesis) and presence (alternate hypothesis) of association of *ACE* I/D with PRF in ADPKD are compared. Third, homogeneity is preferred to heterogeneity, but heterogeneity is unavoidable [46]. Thus, occurrence of heterogeneity between studies was estimated with the χ^2^-based Q test [47], with threshold of significance set at P^b^ < 0.10. Heterogeneity was also quantified with the I^2^ statistic which measures variability between studies [48]. I^*2*^ values of > 50% indicate more variability than those ≤ 50% with 0% indicating zero heterogeneity (homogeneity). Evidence of functional similarities in population features of the studies warranted using the fixed-effects model [49], otherwise the random-effects model [50] was used. Sources of heterogeneity were detected with the Galbraith plot [51] followed by re-analysis (outlier treatment). Of note, outlier treatment dichotomized the comparisons into pre-outlier (PRO) and post-outlier (PSO). Sensitivity analysis, which involves omitting one study at a time and recalculating the pooled OR, was used to test for robustness of the summary effects. We assessed publication bias for comparisons that met two conditions: (i) ≥ 10 studies only [52] and (ii) significant outcomes. Except for heterogeneity estimation [47], two-sided P-values of ≤ 0.05 were considered significant. All associative outcomes were Bonferroni-corrected in order to control for Type 1 error. Data for the meta-analysis were analyzed using Review Manager 5.3 (Cochrane Collaboration, Oxford, England), SIGMASTAT 2.03, and SIGMAPLOT 11.0 (Systat Software, San Jose, CA).

## Results

### Search results and study features

Figure 1 outlines the study selection process in a PRISMA-sanctioned flowchart (Preferred Reporting Items for Systematic Reviews and Meta-Analyses). Initial search resulted in 548 citations, followed by a series of omissions (S1 List) that eventually yielded 18 articles for inclusion [24-41].

**Fig 1.**
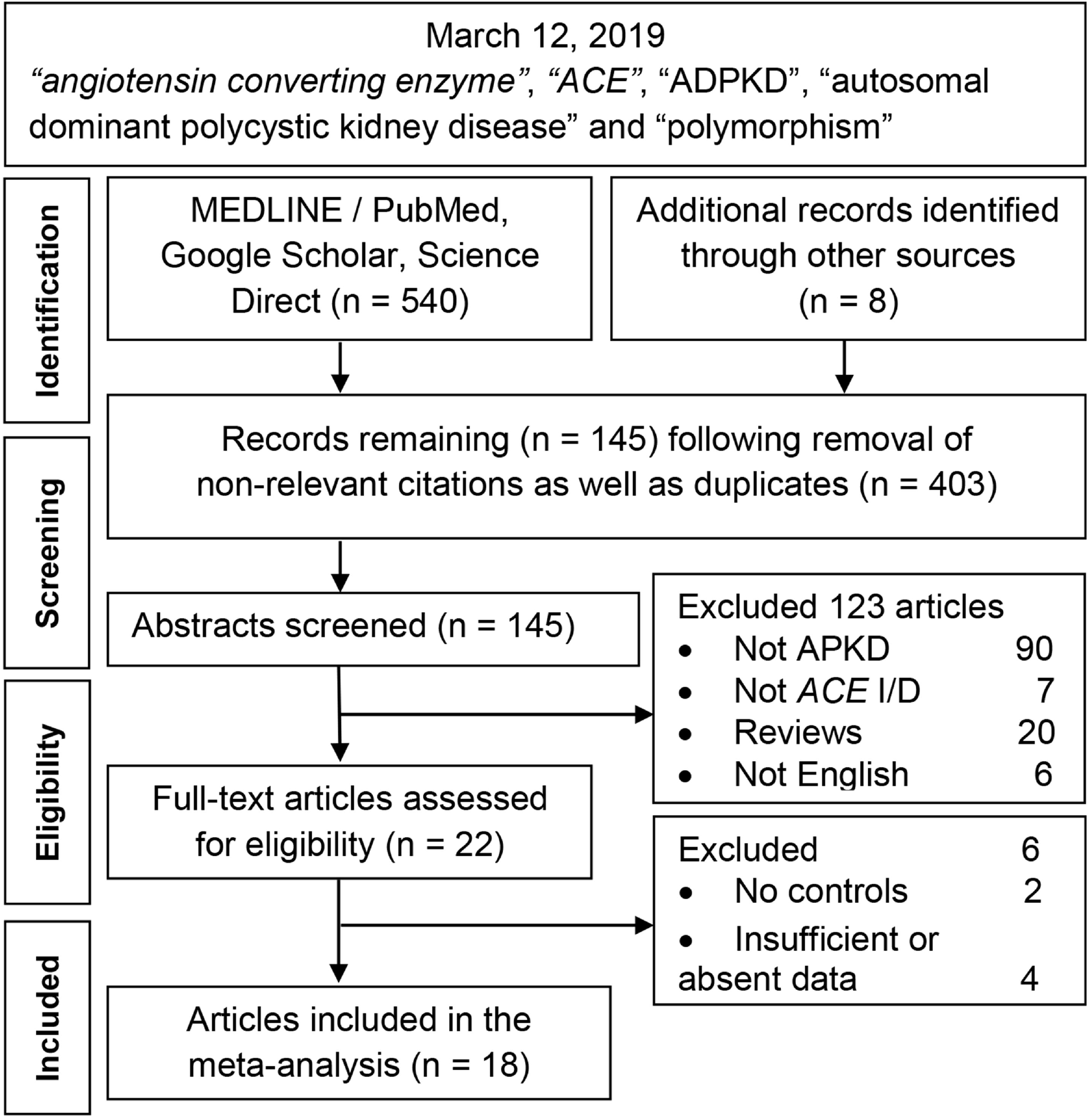
Summary flow chart of literature search.

**S1 List** Excluded studies

### Characteristics of the included studies

Table 1 shows that the number of Caucasian (n = 12) studies was twice that of the Asians (n = 6). Methodological quality of the component studies was moderate based on mean and SD (6.37 ± 1.24) of the normally distributed CB scores (SW test: P = 0.33). S1 Table shows the quantitative features of the included studies, specifically the differential frequencies of the *ACE* genotypes between the race subgroups, where II was higher (83%: 5/6) in the Asian studies and lower (17%: 2/12) in the Caucasian studies. Adequate statistical power was absent in the Asian studies, and present in two Caucasian studies [31,34]. At the aggregate level, however, power in both ethnic subgroups was > 80% (Asian: 97.3%; Caucasian: 100.0%). Control frequencies deviated from the HWE in four studies [27,28,36,41]. This meta-analysis followed the PRISMA and genetic association guidelines (S2-S3 Tables).

### Meta-analysis outcomes

The overall PRO homozygote DD genotype outcome indicating increased risk (OR 1.37) was marginally significant (P^a^ = 0.05) and nullified in the PSO analysis (OR 1.02, P^a^ = 0.78). HWE analysis validated the overall outcomes (Table 2). In the subgroups (Table 3), seven associative outcomes (Asian and ≥ 0.50 maf) were statistically significant, moderate in PRO (P^a^ = 0.01-0.02) and high in PSO (P^a^ < 10^−4^ to 10^−5^). The PSO outcomes survived the Bonferroni-correction, but the PRO did not. The highly significant PSO outcomes had the following features: (i) all were homogeneous (I^2^ = 0%); (ii) magnitude of the homozygote DD increased risk effects was more (ORs 5.01-5.62) than that in heterozygote ID (ORs 1.63-1.68); and (iii) the pooled homozygote II effect indicated reduced risk (OR 0.22). In contrast, Caucasian and < 0.50 maf outcomes were non-associative between *ACE* I/D and PRF in ADPKD (ORs 0.91-1.10, 95% CI 0.79-1.31, P^a^ = 0.15-0.79), and initially non-heterogeneous (fixed-effects) which precluded outlier treatment (Table 3).

**Table 2.**
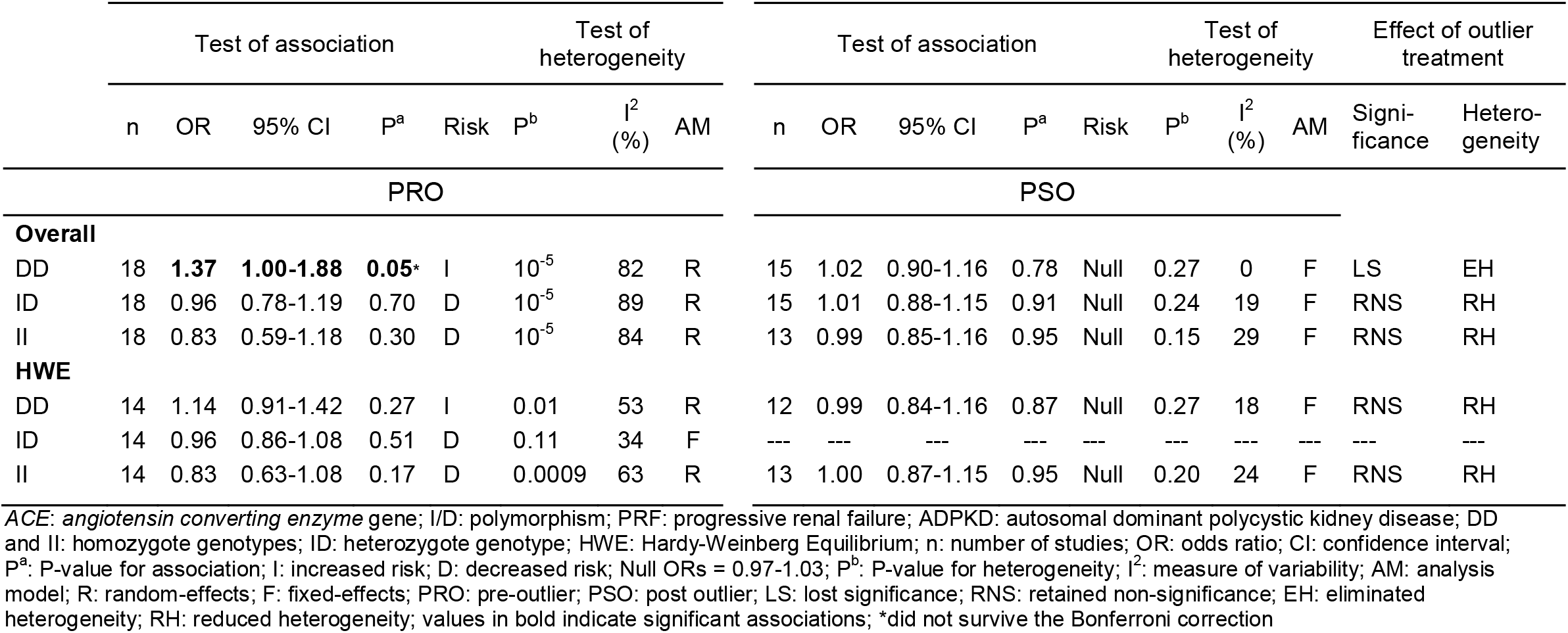
Overall and modified outcomes of *ACE* I/D effects on PRF in ADPKD

**Table 3.**
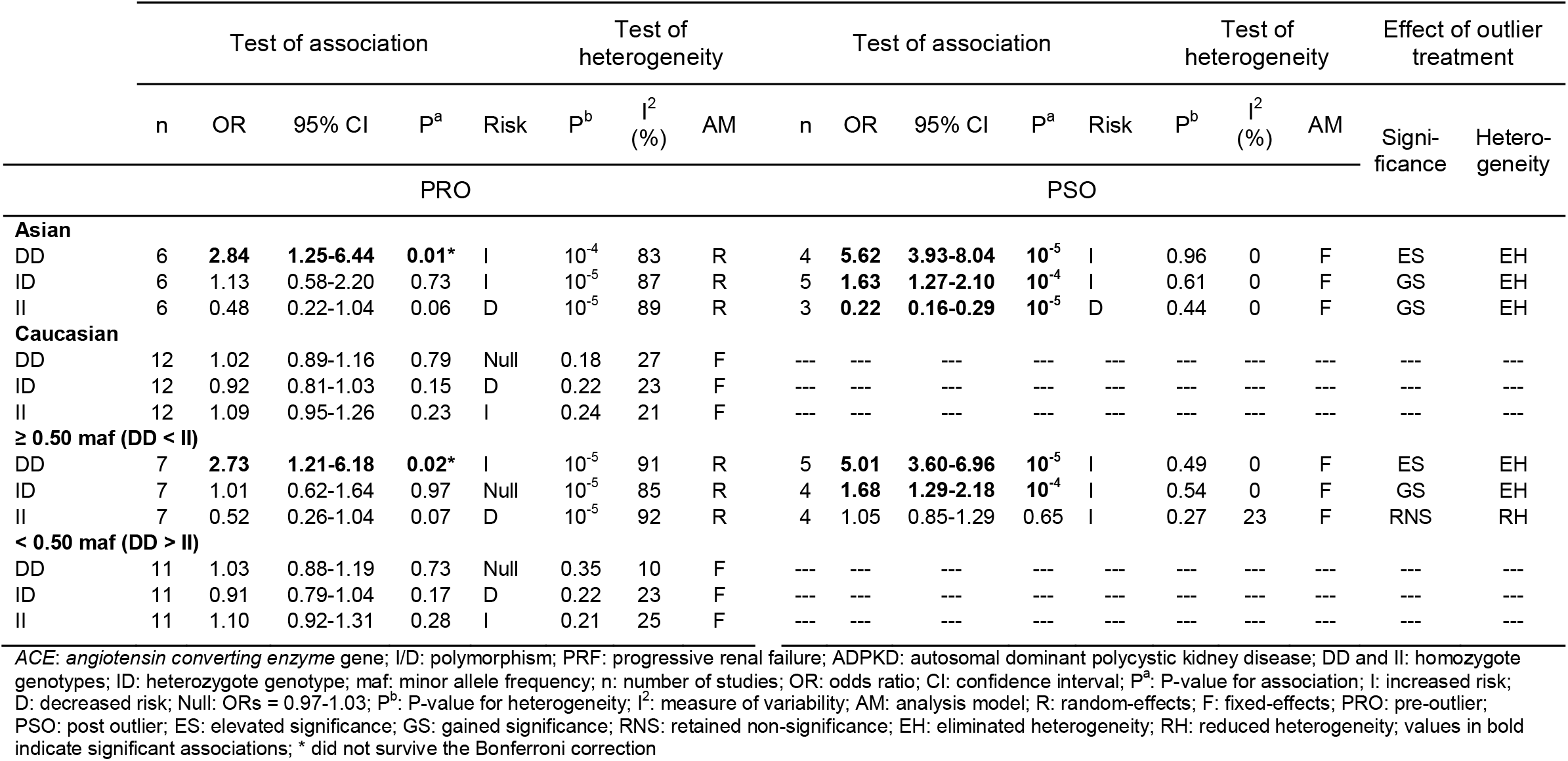
Subgroup effects of the *ACE* I/D polymorphism on PRF in ADPKD

### Mechanism and impact of outlier treatment

The mechanism of outlier treatment for the homozygous DD genotype in the Asian subgroup is visualized in Figs 2-4. Fig 2 shows the PRO forest plot, moderately significant (OR 2.84, 95% CI 1.25-6.44, P^a^ = 0.01) and heterogeneous (P^b^ < 10^−4^, I^2^ = 83%). The Galbraith plot identifies two studies [25,26] as the sources of heterogeneity (outliers), located below the −2 confidence limit (Fig 3). In Fig 4, the PSO outcome (outliers omitted) shows eliminated heterogeneity (P^b^ = 0.96, I^2^ = 0%); intensified increased risk effect (OR 5.62, 95% CI 3.93-8.04) and escalated significance (P^a^ < 10^−5^). This operation is numerically summarized in Table 2.

**Fig 2.**
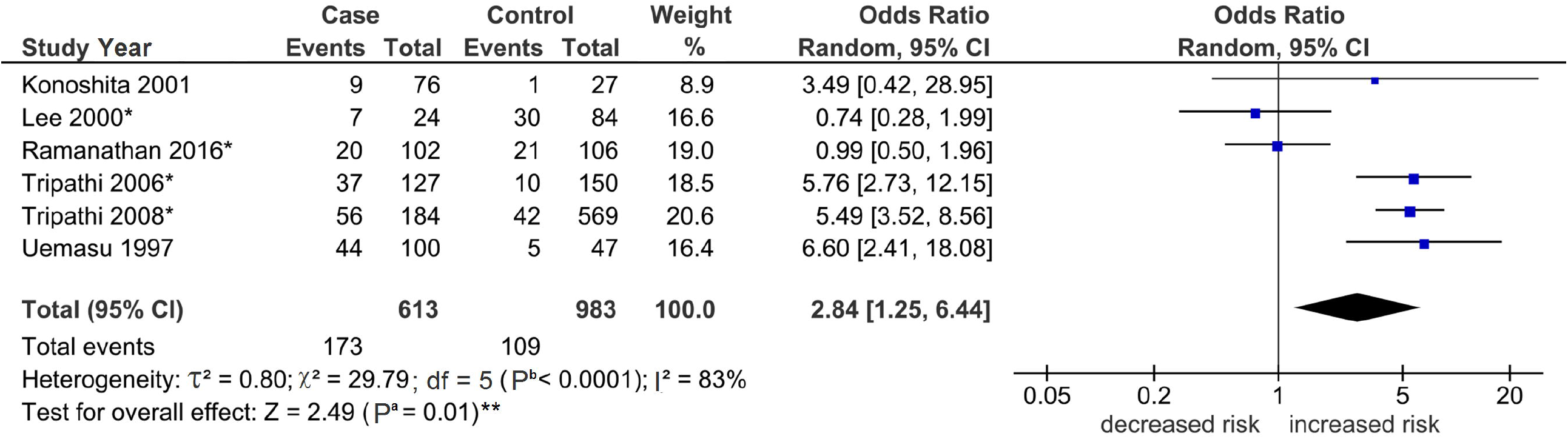
Forest plot outcome of the Asian *ACE* homozygous DD effects on PRF in ADPKD.

**Fig 3.**
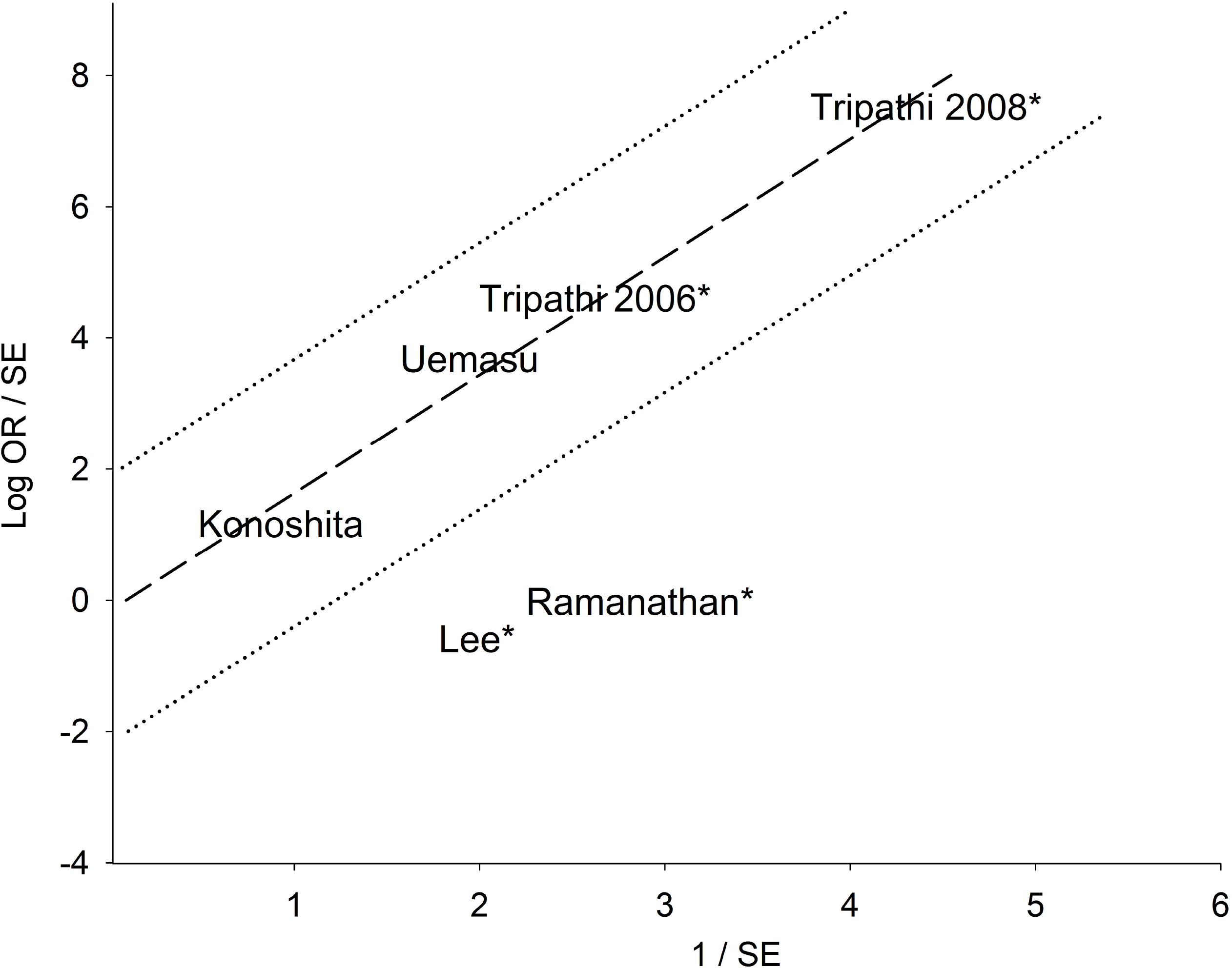
Galbraith plot analysis for the *ACE* homozygous DD genotype in the Asian subgroup.

**Fig 4.**
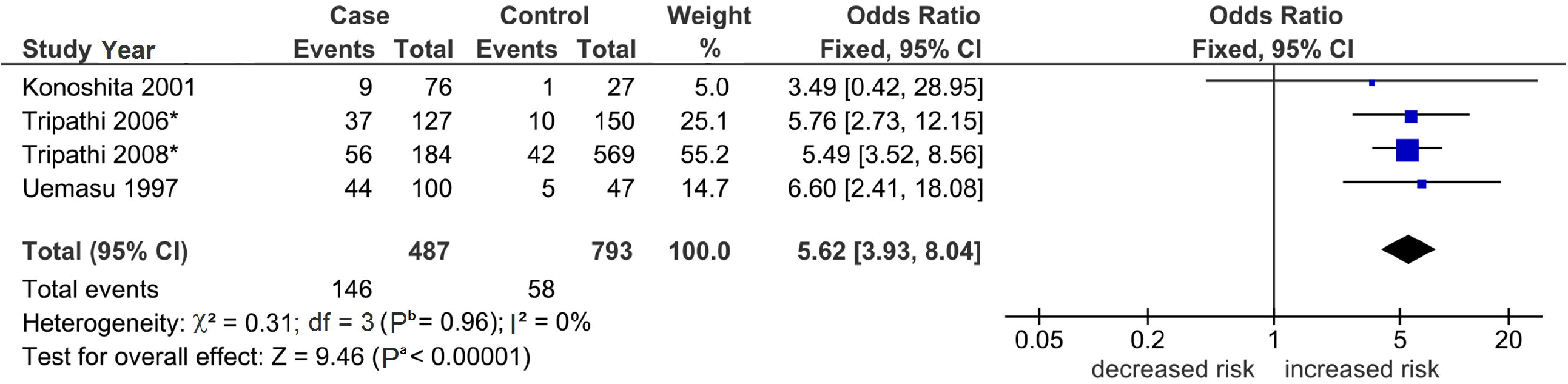
Forest plot outcome of outlier treatment on the Asian *ACE* homozygous DD effects on PRF in ADPKD.

### Sensitivity analysis and publication bias

The high magnitude (ORs 5.01-5.62) of the homozygote DD Asian and ≥ 0.50 maf effects were robust, but not the heterozygote ID and homozygote II genotypes (Table 4). In the DD homozygote genotype, the Asian effects were more robust than the ≥ 0.50 maf subgroup (Table 4). Thus, the Asian DD effects were more stable than that in ≥ 0.50 maf.

**Table 4.** Sensitivity analysis of significant *ACE* I/D ADPKD outcomes

| Comparison | Geno-<br>type | PRO | PSO | A | B |
| --- | --- | --- | --- | --- | --- |
| Overall | DD | Robust | Robust | 0 | 2 |
| Asian | DD | Robust | Robust | 0 | 2 |
| Asian | ID | ----- | [28] | 1 | 0 |
| Asian | II | ----- | [24,26] | 2 | 0 |
| ≥ 0.50 maf | DD | [27,29,38] | Robust | 3 | 1 |
| ≥ 0.50 maf | ID | ----- | [28] | 0 | 1 |
| A |  | 3 | 4 |  |  |
| B |  | 2 | 3 |  |  |

*ACE*: angiotensin converting enzyme gene; I/D: polymorphism; ADPKD: autosomal dominant polycystic kidney disease; maf: minor allele frequency; DD and II: homozygote genotypes; ID: heterozygote genotype; A: Number of references contributing to non-robustness; numbers in brackets indicate references that contributed to non-robustness; B: Number of robust outcomes; PRO: pre-outlier; PSO: post-outlier

Only the PRO overall analysis for the homozygous DD genotype was eligible for publication bias assessment. Non-normal distribution (SW test: P < 10^−4^) of its operating data (ORs) prompted the use of Begg-Mazumdar test of correlation [53] which showed no evidence of publication bias (Kendall’s τ = 0.23, P = 0.19).

### Cumulative meta-analysis

Fig 5 outlines the trend of pooled effects from 1997 to 2016. Here, the “decline effect” is apparent where initial studies show the early extreme phenomenon [54] compared with the subsequent and modulated pooled ORs. Significance was observed in the early studies (1997-1999), lost in the subsequent ones (2000-2006), and then regained in the recent studies (2008-2016). A trend of increasing precision (narrowing of the CIs) is readily observed in the graph.

**Fig 5.**
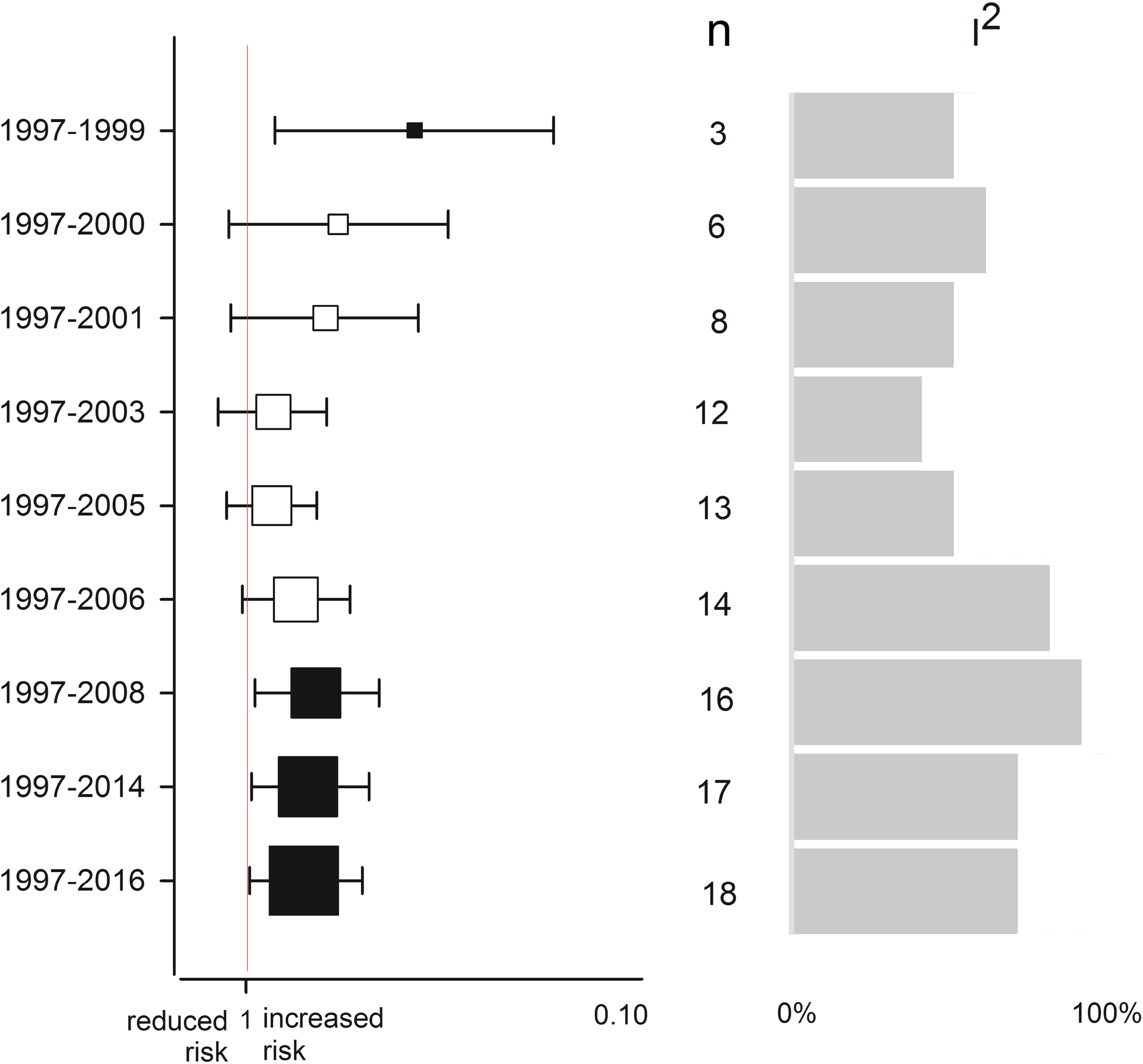
Cumulative meta-analysis of the associations between the *ACE* homozygous DD genotype and PRF in ADPKD.

## Discussion

### Summary of associations

Evidence of association between the *ACE* homozygous DD genotype and risk of PRF in ADPKD is tenuous in the overall analysis but strong in the subgroups (Asian and ≥ 0.50) at the PSO level. This strength rests on the three indicators: (i) high magnitude of effects (up to 5.6-fold), which were (ii) homogeneous (indicating combinability of the studies) and (iii) highly significant (up to P^a^ < 10^−5^). Of note in the Asian subgroup, the II homozygous effect indicating 78% reduced risk also met the high strength of evidence criteria (Table 3), but was found to be non-robust, which contrasted with robustness of the homozygous DD effect. Our use of subgroup analysis and outlier treatment have unraveled significant and homogeneous associations in three contexts, (i) not achieved in a previous meta-analysis; (ii) neither present in the overall analysis; (iii) nor in the component single-study outcomes. However, the process of subgrouping and performing outlier treatment reduced the number of studies and sample sizes of the comparisons. These may decreased statistical power and risked Type 1 error, especially in light of a few (three to five) PSO studies generating multiple significant outcomes in the subgroup analyses. Fortunately, adequate statistical powers of the three to five Asian studies (82.2-97.3%) and their P-values surviving the Bonferroni correction served to minimize the possibility of false-positives and increase confidence in the outcomes. Non-associative outcomes in previous primary studies may be attributed to their lack of power and small sample sizes. Underpowered outcomes appear to be common in candidate gene studies [55] and are prone to the risk of Type 1 error. Other highlights of our findings are differential effects between the ethnic (Asian versus Caucasian) and maf (≥ 0.50 versus < 0.50) subgroups. Significant outcomes in Asian but not in Caucasians may be attributed to differences in maf between the two ethnicities [56].

### Cumulative meta-analysis

Studies on associations between *ACE* I/D and PRF in ADPKD from the last 20 years produced outcomes that reduced the magnitude and retained the heterogeneity of associations. On the other hand, the sustained increased precision and cumulative significance of effects as well as increase in the number of studies may help establish evidence of association.

### Comparison with other meta-analyses

Five previous meta-analyses relate to our study, four of them not as direct as the fifth. The first [57] focused on angiotensin II types 1 and 2 receptor genes (*AGTR1* and *AGTR2*) in various renal diseases that did not include ADPKD. The second to the fourth meta-analyses [13,14,58] focused on *ACE* I/D with the outcome of end-stage renal disease (ESRD). All three demonstrated associations of *ACE* I/D with ESRD, two of which drew race-specific conclusions, one for Caucasians [58] and the other for Asians [14]. The fifth [21] directly relates to our study which was published 13 years ago (2006); the features of both are outlined in Table 4. Pereira et al [21] included non-English articles and meeting abstracts, which we did not in our study because we evaluated methodological quality of the included studies. Methodology-wise, Pereira et al used the recessive model (homozygous DD versus heterozygous ID + homozygous II genotypes) versus the genotype model (homozygous DD, and II, heterozygous ID) in ours. Comparative outcomes are based on the homozygous DD genotype (Table 5). Three differences mark the overall analyses (ours and the previous): (i) the number of studies (18 versus eight); (ii) significance of outcome (P^a^ = 0.05 versus P = 0.21) and (iii) magnitude of outcome (1.4-fold versus 1.2-fold). In the racial subgroups, Pereira et al showed no material differences between the Asian and Caucasian outcomes (both indicated non-significant increased risks, randomly-derived). In contrast, our outcomes differentiated between the Asians and Caucasians (up to 6-fold versus null). In the Asian outcomes, ours and the previous differed in terms of: (i) significance (P^a^ = 0.01-10^−4^ versus P = 0.23) and magnitude of effect (up to 5.6-fold versus 1.6-fold). In the Caucasian analyses, ours versus previous differed in the number of studies (12 versus four) and outcomes (null versus 1.2-fold).

**Table 4.** Comparison of features between the two meta-analyses

|  | This study | Pereira et al [21] |
| --- | --- | --- |
| **General features** |  |  |
| Year | 2019 | 2006 |
| Country | Thailand | Brazil |
| Subgroups | Race/minor allele frequency | Race |
| Genetic modeling | Allele/Genotype | Recessive |
| **Methods / Treatments** |  |  |
| HWE as inclusion criterion | Yes | No |
| Modifier analysis | Yes | No |
| Outlier analysis | Yes | No |
| Heterogeneity | Q statistic, $I^2$ | Q statistic |
| Sensitivity analysis | Yes | No |
| Publication bias | Yes | Yes |

HWE: Hardy-Weinberg Equilibrium

**Table 5.**
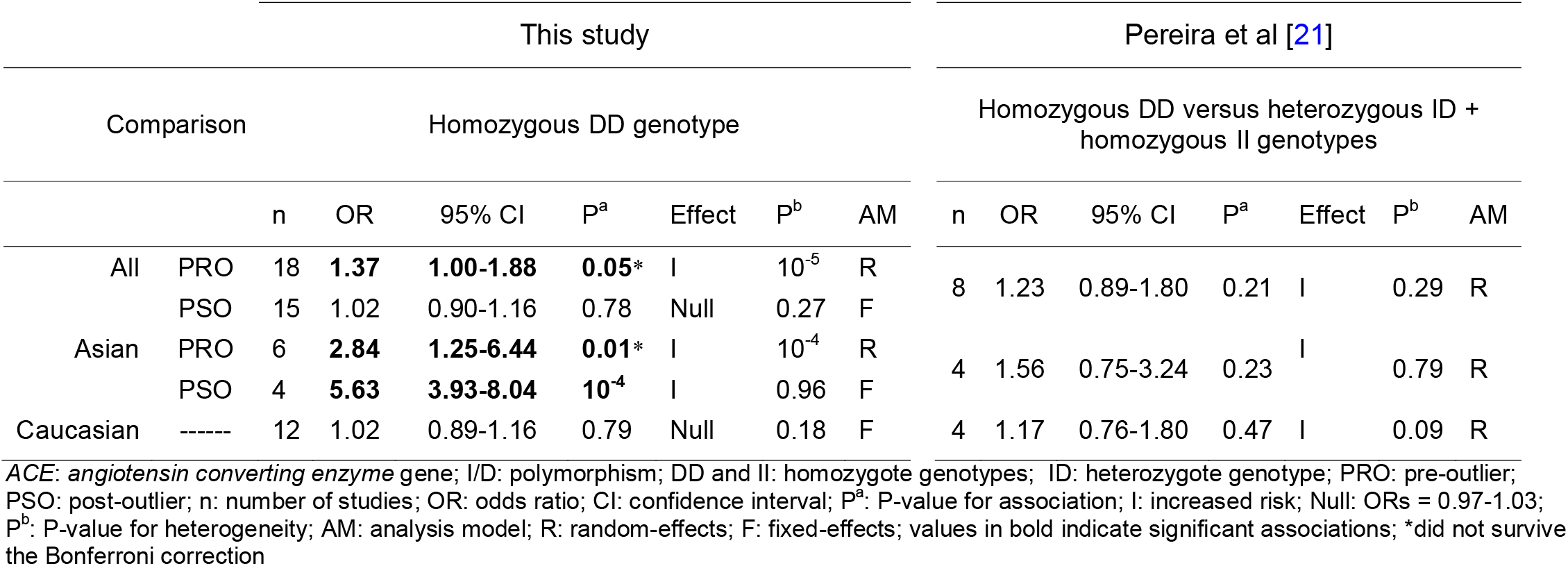
Comparison of outcomes between the current meta-analysis and the 2006 meta-analysis

|  |  | This study |  |  |  |  |  |  | Pereira et al [21] |  |  |  |  |  |  |
| --- | --- | --- | --- | --- | --- | --- | --- | --- | --- | --- | --- | --- | --- | --- | --- |
| Comparison |  | Homozygous DD genotype |  |  |  |  |  |  | Homozygous DD versus heterozygous ID + homozygous II genotypes |  |  |  |  |  |  |
|  |  | n | OR | 95% CI | P<sup>a</sup> | Effect | P<sup>b</sup> | AM | n | OR | 95% CI | P<sup>a</sup> | Effect | P<sup>b</sup> | AM |
| All | PRO | 18 | **1.37** | **1.00-1.88** | **0.05\*** | I | 10<sup>-5</sup> | R | 8 | 1.23 | 0.89-1.80 | 0.21 | I | 0.29 | R |
|  | PSO | 15 | 1.02 | 0.90-1.16 | 0.78 | Null | 0.27 | F |  |  |  |  |  |  |  |
| Asian | PRO | 6 | **2.84** | **1.25-6.44** | **0.01\*** | I | 10<sup>-4</sup> | R | 4 | 1.56 | 0.75-3.24 | 0.23 | I | 0.79 | R |
|  | PSO | 4 | **5.63** | **3.93-8.04** | **10<sup>-4</sup>** | I | 0.96 | F |  |  |  |  |  |  |  |
| Caucasian | ----- | 12 | 1.02 | 0.89-1.16 | 0.79 | Null | 0.18 | F | 4 | 1.17 | 0.76-1.80 | 0.47 | I | 0.09 | R |

ACE: *angiotensin converting enzyme* gene; I/D: polymorphism; DD and II: homozygote genotypes; ID: heterozygote genotype; PRO: pre-outlier; PSO: post-outlier; n: number of studies; OR: odds ratio; CI: confidence interval; P<sup>a</sup>: P-value for association; I: increased risk; Null: ORs = 0.97-1.03; P<sup>b</sup>: P-value for heterogeneity; AM: analysis model; R: random-effects; F: fixed-effects; values in bold indicate significant associations; \*did not survive the Bonferroni correction

### Genetic correlates

Understanding the impact of *ACE* I/D on renal disease might clarify the role of the homozygous DD genotype in PRF and/or variable responses to renoprotective therapy. Based on the evidence that the homozygous DD genotype elicits high ACE activity [7], it was hypothesized that the homozygous DD genotype were resistant to renoprotective therapy [9]. However, it has also been hypothesized that *ACE* I/D is in linkage disequilibrium with an unknown DNA section containing a silencer motif that could inhibit ACE mRNA translation [59,60]. This suggests variability of ACE expression at the DNA/mRNA level. Studies have reported increased ACE mRNA expression in homozygous DD genotype patients in renal [61] and other tissues [62,63]. It is then possible to identify different polymorphisms in the *ACE* gene to identify patients at risk for PRF or therapy resistance. Evidence for the association between serum ACE levels and ADPKD has not been consistent [33,39]. The numerous studies that examined associations at the gene level had conflicting outcomes. An association between the D allele of *ACE* I/D among ADPKD patients has been detected in various populations [24,30,37,40], but not in others [25,29,32,33,35,36,39].

### *ACE* gene, ACE protein and RAAS

The renin-angiotensin aldosterone system (RAAS) is critical in regulating blood pressure and electrolyte balance [64,65]. The activity of RAAS system in turn, is regulated by the ACE protein where it mediates the rate of production of angiotensinogen from renin [66]. RAAS is activated in progressive kidney disease leading to hypertension [67]. The role of hypertension in the progression of kidney deterioration in ADPKD places the *ACE* I/D polymorphism at the core of this research. On a morphological level, the pathophysiology of ADPKD involves RAAS where it directly stimulates the growth of renal cysts [68]. Angiotensin II, a RAAS component was observed within cysts and tubules leading to the cyst expansion [69].

### Strengths and limitations

Interpreting our findings should consider its limitations and strengths. Limitations include: (i) Eighty-nine percent of the component studies were underpowered; (ii) of the 29 comparisons, 22 (76%) were heterogeneous; (iii) pathological conditions of the some cases were diluted, where ADPKD comprised part of the patients’ condition and (iv) despite the high significance (P^a^ < 10^−5^) and magnitude (5-fold) as well as homogeneity (I^2^ = 0%) of the homozygous DD PSO pooled effects in the Asian and ≥ 0.50 maf comparisons, their wide CIs (3.60-8.04), suggests imprecision which may have compromised the durability of effects. Larger sample sizes may be needed to ameliorate this limitation [70]. Of note, precision in this study had opposing effects, reduced (wide CIs) with subgroup treatment but increased (narrow CIs) with cumulative meta-analysis. The methodology may explain this paradox in terms of sample size and number of studies (n), which was reduced with subgroup analysis but increased with cumulative meta-analysis.

On the other hand, the strengths comprise of the following: (i) the combined sample sizes of the overall and subgroups translated to high statistical power; (ii) ten (56%) of the 18 articles addressed HWE concerns. Confining the analysis to studies in HWE did not materially alter the pooled ORs; in fact, HWE-analysis validated the overall pooled effects. Given this outcome, the risk of genotyping errors appears to be a minor issue which minimizes methodological weakness in our study; (iii) outlier treatment reduced and eliminated heterogeneity and enabled significance; (iv) the significant PSO ORs in the Asian and ≥ 0.50 maf subgroups survived the Bonferroni correction, thus minimizing the possibility of a Type 1 error; (v) sensitivity treatment conferred robustness to the overall and Asian homozygous DD findings and (vi) publication bias was not evident in the PRO overall comparison.

## Conclusions

We have shown that associations of *ACE* I/D with PRF in ADPKD are genotype dependent. Substantial amount of evidence presented here may render *ACE* I/D useful as prognostic markers for PRF in ADPKD. In spite of the evidence for associations, the complexity of ADPKD involves interactions between genetic and non-genetic factors allowing for the possibility of environmental involvement. Gene-gene and gene-environment interactions have been reported to have roles in associations of other polymorphisms with ADPKD. All but five [29,32,38,39,41] of the 18 articles acknowledged gene-environment interaction.

Addressing gene-gene and gene-environment interactions [71] may help address the pathophysiological significance of *ACE* I/D and PRF in ADPKD. Four of the included articles mentioned haplotype analysis [26,27,35,39] with one presenting haplotype data [26]. Focus on *ACE* haplotypes have been suggested for future association studies [72]. Additional well-designed studies exploring other parameters would confirm or modify our results in this study and add to the extant knowledge about the association of the *ACE* polymorphism and PRF in ADPKD.

## Data Availability

All relevant data are within the paper and its Supporting information files

## List of abbreviations

ABP: Australia Bulgaria Poland
*ACE*: angiotensin converting enzyme gene
ACE: angiotensin converting enzyme protein
ADPKD: autosomal dominant polycystic kidney disease
AM: analysis model
BF: Bayes Factor
BFR: Belgium France
CB: Clark-Baudouin
CC: case-control
CI: confidence interval
CO: cohort
CS: cross-sectional
D: decreased risk
DD: variant homozygous genotype
DNA: deoxyribonucleic acid
EH: eliminated heterogeneity
ES: elevated significance
F: fixed-effects
GS: gained significance
HWE: Hardy-Weinberg Equilibrium
I: increased risk
I^2^: measure of variability
I/D: polymorphism
ID: heterozygous genotype
II: common homozygous genotype
LS: lost significance
maf: minor allele frequency
mRNA: messenger ribonucleic acid
N: total number of comparisons
n: number of studies
*n*: frequency of occurrence
OR: odds ratio
P^a^: P-value for association
P^b^: P-value for heterogeneity
PRF: progressive renal failure
PRISMA: Preferred Reporting Items for Systematic Reviews and Meta-Analyses
PRO: pre-outlier
PSO: post-outlier
R: random-effects
[R]: Reference of studies
RNS: retained non-significance
RH: reduced heterogeneity
UKA: United Kingdom Australia
USA: United States of America

## Author contributions

**Conceptualization:** NP, PT

**Data curation:** NP, PT and YP

**Formal analysis:** NP, PT and YP

**Investigation:** NP, PT

**Methodology:** NP, PT and YP

**Project administration:** NP, PT

**Resources:** PT, NP and AT

**Software:** NP, PT

**Supervision:** NP

**Validation:** NP, PT, YP and AT

**Visualization:** NP, PT and YP

**Writing – original draft:** NP, PT

**Writing – review & editing:** NP, PT, YP and AT

## Data availability statement

All relevant data are within the paper and its Supporting information files

## Supporting information

S1 List Excluded articles DOCX

S1 Table Quantitative DOCX

S2 Table PRISMA checklist DOCX

S3 Table Genetic checklist DOCX

S5 Data Raw data code XLS

---

**S1 Table** Quantitative features of the included studies

**S2 Table** PRISMA checklist

**S3 Table** Meta-analysis checklist

